# Automated method to extract and purify RNA from wastewater enables more sensitive detection of SARS-CoV-2 markers in community sewersheds

**DOI:** 10.1101/2022.04.03.22273370

**Authors:** Nicholas W. West, Adrian A. Vasquez, Azadeh Bahmani, Mohammed Khan, James Hartrick, Carrie L. Turner, William Shuster, Jeffrey L. Ram

## Abstract

Wastewater based epidemiology (WBE) has emerged as a strategy to identify, locate, and manage outbreaks of COVID19, and thereby possibly prevent surges in cases, which overwhelm local to global health care networks. The WBE process is based on assaying municipal wastewater for molecular markers of the SARS-CoV-2 virus. The standard process for sampling municipal wastewater is both time-consuming and requires the handling of large quantities of wastewater, which negatively affect throughput and timely reporting, and can increase safety risks. We report on a method to assay multiple sub-samples of a bulk wastewater sample. We document the effectiveness of this new approach by way of comparison of technologies for automating RNA purification from wastewater samples. We compared processes using the Perkin-Elmer Chemagic™ 360 to a PEG/NaCl/Qiagen protocol that is used for detection of N1 and N2 SARS-CoV-2 markers by the majority of 19 pandemic wastewater testing labs in the State of Michigan. Specifically, we found that the Chemagic™ 360 lowered handling time, decreased the amount of wastewater required by 10-fold, increased the amount of RNA isolated per µl of final elution product by approximately five-fold, and had no deleterious effect on subsequent ddPCR analysis. Moreover, for detection of markers on the borderline of detectability, we found that use of the Chemagic™ 360 enabled the detection of viral markers in a significant number of samples for which the result with the PEG/NaCl/Qiagen method was below the level of detectability. This improvement in detectability of the viral markers might be particularly important for early warning to public health authorities at the beginning of an outbreak.

## Introduction

The rapid global spread of severe acute respiratory syndrome coronavirus 2 (SARS-CoV-2), the RNA virus that causes coronavirus disease 2019 (COVID-19), has put enormous strain on healthcare institutions and on epidemiological efforts to monitor, track, and predict the spread and evolution of SARS-CoV-2. Early knowledge of the sequences of the SARS-CoV-2 virus (Wu et al., 2020) enabled molecular tests for the presence of the virus to be developed for its presence in both clinical samples and in virus shed by infected individuals into wastewater. Community-wide monitoring efforts in the United States were hampered initially by applying tests only to symptomatic individuals who had traveled from China and by lack of availability of tests (Bendix, 2020; CDC Health Alert Network, 2020). However, even with tests subsequently made more widely available, resistance or low response rates to requests to be tested have occurred in various settings, including college campuses (Gibas et al., 2021). The testing of only symptomatic individuals was especially problematic for controlling the disease because a delay occurs between infection and capability to spread the virus and the occurrence of symptoms. Furthermore, some individuals remain asymptomatic even with observed viral RNA quantities similar to those presenting symptoms (Arons et al., 2020; Oran and Topol, 2021).

For assessing community levels of COVID-19 disease, detection of the virus in wastewater can be a successful strategy. Molecular markers for SARS-CoV-2 virus are shed in feces early in infection and have been observed in raw wastewater collected from locations where infected people are present (Ahmed et al., 2020). Wastewater monitoring has been used to surveil diseases and other public health relevant markers in the past (Castiglioni et al., 2006; Heijnen and Medema, 2011; Brouwer et al., 2018; Xiao et al., 2019). Accordingly, wastewater based epidemiology of SARS-CoV-2 has been underway in multiple locations (Naughton, 2020; Rose et al., 2021). The State of Michigan funded 19 health department and university laboratories across the state to implement wastewater analysis of SARS-CoV-2 for the purposes of providing early warning signs of COVID-19 infections in various communities across the state (State of Michigan, 2021).

Numerous reports have confirmed the value of detecting SARS-CoV-2 markers in wastewater for public health purposes. Researchers have found that wastewater measurements of SARS-CoV-2 markers provide population-level insights including data about disease prevalence and predicted symptomatic cases and hospitalizations (Panchal et al., 2021; Galani et al., 2022). Wastewater monitoring detected SARS-CoV-2 six to eight days before positive tests were reported in New Haven, Connecticut, USA (Peccia et al., 2020), 12 - 16 days before COVID-19 cases were declared in four municipalities in Spain (Randazzo et al., 2020) and before the exponential growth of the epidemic in Paris, France (Wurtzer et al., 2020). A further advantage of measuring SARS-CoV-2 markers in wastewater as a way of assessing the presence of COVID-19 in the community is that such tests can be accomplished with a comparatively small burden on local healthcare resources compared to frequent and intensive testing of multiple individuals.

In order to successfully detect viral particles from large volumes of wastewater from hundreds to thousands of individuals, the viral RNA in the wastewater samples must first be concentrated and purified. The two steps are usually accomplished sequentially and listed in separate columns of methods review tables (see, for example, Ali et al. (2021) and Alygizakis et al. (2021)). Among previously used methods for concentrating RNA viruses from wastewater are precipitation with polyethylene glycol (PEG) 8000 and high salt followed by centrifugation (Ye et al., 2016; Borchardt et al., 2017), ultracentrifugation (Ye et al., 2016), and ultrafiltration (Borchardt et al., 2017; Medema et al., 2020). A recent study by Flood et al. (2021) compared all three methods using spiked-in *Pseudomonas* phage Phi6 to compare efficiency of these methods in concentrating enveloped RNA viruses from wastewater. The PEG method significantly increased recovery of Phi6, yielding between 2-times and up to 13-times more Phi6 detected in PEG concentrate than the in the concentrate isolated by ultracentrifugation or ultrafiltration methods. However, as noted above, concentration is usually followed by a subsequent purification step. For example, in the study by Flood et al. (2021), RNA from wastewater was purified from the concentrate on silica-based spin columns (QIAmp Viral RNA Minikit by Qiagen). Other purification methods have been based on binding and selective elution from magnetic silica beads (Biomerieux Nuclisens kit), as implemented by Medema et al. (2020) and many others (Ali et al., 2021).

These two step procedures (concentration first, then purification) are time-consuming and may be equipment-intensive. For example, the PEG/NaCl/Qiagen protocol recommended for use in the State of Michigan is based on the method described by Flood et al. (2021) and comprises several hours precipitation time and centrifugation of large volumes (100 mL) of wastewater prior to removal of supernatant, transfer of pellets that may vary in size and consistency, and ultimately running the concentrated extract through Qiagen columns. The method may also allow PCR inhibitors to accompany the final product resulting in varying analytical efficiency (Monteiro et al., 2022). In the present study, we sought to combine the concentration and purification steps into a single automated procedure using a large-volume approach based on magnetic bead technology.

We describe here our success in achieving improved recovery of RNA with low levels of PCR inhibitors based upon using a rapid automated method on the Perkin-Elmer Chemagic™ 360 platform. During our preliminary investigations prior to implementation of the Chemagic™ 360 system in our laboratory, we also explored the potential use of several other automated systems (specifically, those by ThermoFisher and by Promega). The Discussion section of this paper provides additional comparative considerations (time required, number of manual or preliminary steps, cost of reagents, cost of equipment, etc.) that might be considered in choosing an improved method for purification of viral RNA from wastewater.

## Methods

### Collection of wastewater

Wastewater was collected once a week from 20 different sites in the Detroit, MI. Access to the sewer system at each site was by manhole, with many being located in local roads. Samples for the present study were collected between 7 am and 9:30 am on each collection day. Each sample consisted of a grab sample of approximately 250 ml wastewater collected from each site using a 250-ml or 500-ml high density polyethylene bottle. At least one out of every 10 samples was collected as a “field blank” of deionized water transferred in the field into a high density polyethylene bottle identical to those used for wastewater samples. Sample bottles were placed in a Ziplock bag with a sheet of paper towel and stored on ice at 4 - 6 °C in coolers until delivery to the laboratory by 10:15 a.m.

Chain-of-custody (CoC) forms were used to document sample collection information (location, date-time of sample collection, sample type [grab vs. composite] and record the transfer of sample custody from the field sampling crew to the laboratory. Physical parameter data, including pH, temperature, dissolved oxygen, and specific conductance, were also measured at each sampling location on each day of sampling using a YSI ProDSS with GPS (YSI, Yellow Springs, OH). The data is collected on a physical field sheet that is transferred to the lab after collection. A digital record of each CoC and each YSI data sheet has been preserved and archived with the project records.

### PEG/NaCl/Qiagen method to assay wastewater

This study directly compared the Chemagic™ 360 method to the PEG/NaCl/Qiagen method, as described by Flood et al. (2021) and in protocols provided by the State of Michigan. The PEG/NaCl/Qiagen method was required to be used by our lab by our funding agency (Michigan Department of Health and Human Services) until we were able to demonstrate the improved results obtained with the Chemagic™ 360 system. A brief description of the PEG/NaCl/Qiagen method is as follows: 100 mL of wastewater was added to 8 g PEG and 1.7 g NaCl, mixed well, held at 4 °C for 2 hrs, then centrifuged at 4696 x g at 4 °C for 45 min. Supernatant was removed, leaving a soft pellet of 2 to 4 mL volume (depending on the sample quality and non-fecal matter). RNA in 200 µl of the pellet was then purified on Qiagen spin columns, yielding 80 µL of purified RNA. Depending on the purpose of the experiment, *Phi6* virus was added as an internal standard either to the 100 mL of wastewater (10 µL of 10^8^ PFU/mL added to 100 mL of wastewater) prior to the addition of PEG and NaCl) or, alternatively, to the 200 µl pellet aliquot (10^6^ PFU/µL of pellet) prior to purification on the Qiagen column.

### Chemagic™ 360 method

We used a Chemagic™ 360 instrument (catalog number, 2024-0020) with the 12-rod head (CMG-371, PerkinElmer Health Sciences Inc., Shelton, CT, USA), capable of processing 10 mL wastewater samples without prior concentration. Previous publications have described use of the Chemagic™ instrument with a 96-rod head, capable of processing 1 mL samples, for which RNA in the wastewater sample must first be concentrated from a larger volume by a time-consuming concentration step such as PEG/NaCl (Laturner et al., 2021) or ultracentrifugation (Hokajärvi et al., 2021); however, this is the first analysis of the Chemagic™ instrument used to concentrate and purify PCR-ready viral RNA from 10 mL wastewater samples in a single integrated Chemagic™-based procedure without a preceding concentration step required. The method is described in detail in a published protocol (Vasquez et al., 2021) and briefly here: Prior to placing samples on the automated instrument, 45 mL of the wastewater sample is centrifuged at 4,696 x g; 10 mL of the supernatant is transferred and mixed into a tube containing Poly A RNA (7 µL Perkin-Elmer CMG842), Proteinase K (50 µL Perkin-Elmer CMG749), and lysis buffer 1 (8 mL Perkin-Elmer CMG749); incubated for 30 min at 55 °C; and then magnetic beads (50 µL Perkin-Elmer CMG749) are added and mixed. After placement of the incubated sample into the Chemagic™ 360 instrument and a set of receiving tubes (4 mL Sarstedt®, containing 100 µL elution buffer CMG749), the instrument is run with protocol *chemagic™Viral10k 360 H12 prefilling drying VD210119*.*che* for 75 min. The product, in the Sarstedt tube in a final volume of ∼ 85 µL elution buffer, is then transferred to a 1.5 mL Lo-Bind centrifuge tube for subsequent analysis or long-term storage at -80 °C. Depending on the purpose of the experiment, *Phi6* virus was added as an internal standard either to the 10 mL of wastewater (10 µL of 10^6^ PFU/mL added to 10 mL wastewater) after the initial centrifugation or, alternatively, to the eluted sample (10 µL of 10^6^ PFU/µL to the eluate) or to elution buffer that had not undergone processing (reference positive control; 10 µL 10^6^ PFU/mL added to 75 µL elution buffer).

### ddPCR Analysis

Primers and TaqMan® probes were designed to amplify and detect nucleocapsid gene markers 1 and 2 (N1 and N2) of SARS-CoV-2 and the P8 nucleocapsid gene of bacteriophage Phi6. The primer and probe sequences used in this study are shown in Table 1.

**Table 1:**
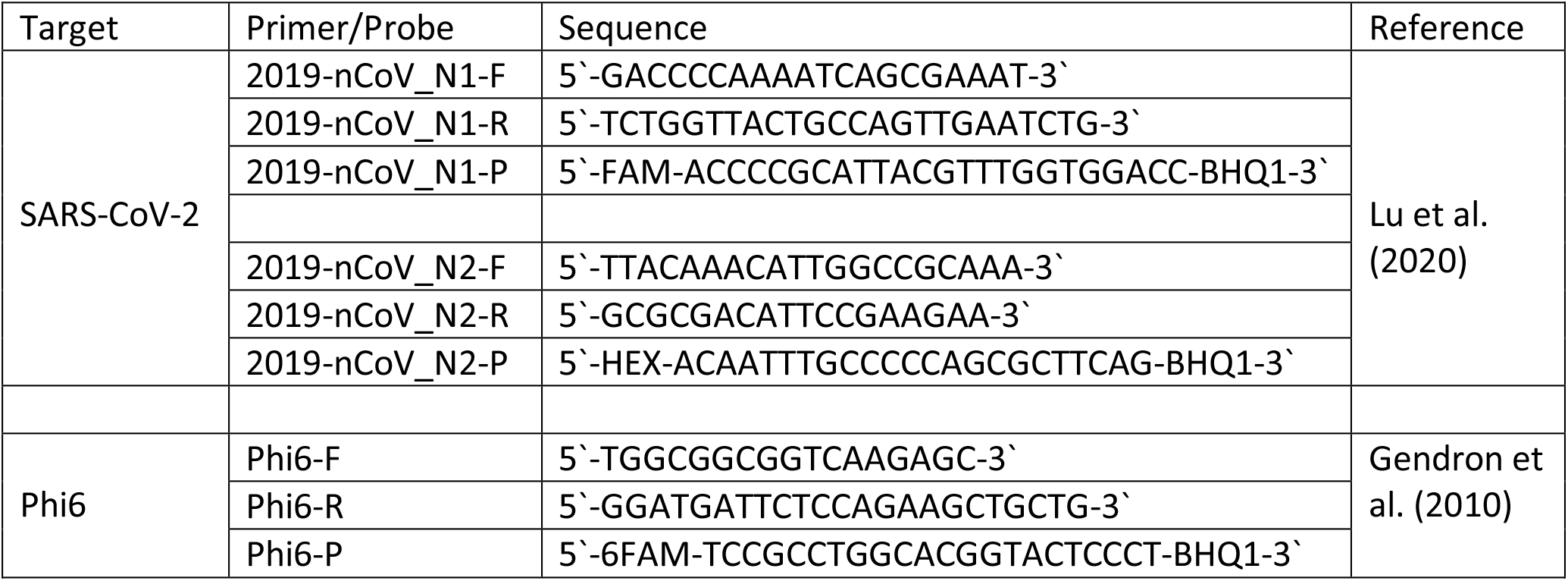
Primer and probe sequences used for ddPCR

Reactions are assembled using Bio-Rad’s One-step RT-ddPCR Advanced Kit for Probes and their suggested protocol (Bio-Rad, CA, USA). Briefly, a 16.5 µL reaction is prepared to contain a final concentration of 1 x Supermix, 20 U/µL reverse transcriptase, 15 mM DTT, 900 nmol/µL of gene target primers, 250 nmol/µL of gene target probe, and 1.1 µL RNAse-free water. TaqMan® probes are labeled with different fluorescent markers enabling primers and probes for both N1 and N2 to be included in the same reaction with duplex detection. To this reaction 5.5 µL of template nucleic acids purified either by Chemagic™ or PEG/NaCl/Qiagen is added for a final reaction mix of 22 µL, of which 20 µL is processed for each ddPCR reaction. ddPCR reactions for each sampling site and quality controls (positive, no-template, extraction, and processing controls) are prepared in triplicate and loaded onto a 96-well PCR plate. The plate is sealed using the Bio Rad PX1 PCR Plate Sealer at 180 °C for 5 sec. The plate is vortexed thoroughly and centrifuged for 1 min at 850 x g on a microplate centrifuge (Thermo Fisher Scientific, MA, USA) and then transferred to the QX200 Automated Droplet Generator (Bio-Rad, CA, USA). Upon completion of droplet generation according to the manufacturer’s instruction, the 96-well plate containing droplets is removed and sealed with the PX1 PCR Plate Sealer at 180 °C for 5 sec.

The plate is then transferred to the Bio-Rad C1000 Thermo Cycler and PCR is initiated with the following settings: Hold 25 °C for 3 min., reverse transcription 50 °C for 60 min., enzyme activation 95 °C for 10 min, denaturation 95 °C for 30 sec., annealing/extension 55 °C for 1 min, denaturation and annealing/extension steps cycled 40 times, enzyme deactivation 98 °C for 10 min, and hold 4 °C until ready for droplet reading. The 96-well plate is transferred to the Bio-Rad QX200 Droplet Reader which is prepared for analysis by using the Bio-Rad QuantaSoft software package version 1.7.4.0917. Once droplet reading is complete and signal threshold values entered for each reaction the resulting file is exported in comma separated value (.csv) format for subsequent analysis.

Results were calculated in terms of # of copies of PCR targets per 100 mL of the original wastewater (or field blank) sample as described by Flood et al. (2021) for PEG/NaCl/Qiagen. Similarly, adjustment of various volumes in the calculation enabled a comparable calculation of SARS-CoV-2 per 100 mL wastewater for samples processed here by Chemagic™ 360.

## Results

### Recovery of spiked-in Phi6

#### Amount of spiked-in Phi6 purified from field blank compared to eluate blank

To assess the ability of the Chemagic™ method to recover a known amount of Phi6 without any interfering factors from wastewater, we compared the ddPCR copies detected from Phi6 spiked into 10 mL of pure water (field blank control) to the same amount of Phi6 spiked directly into elution buffer (eluate control) similar in volume to the final eluate from the Chemagic™ procedure. For sixteen pairs of spiked-in field blank and eluate controls analyzed between 15 November 2021 and 29 December 2021, the Phi6 measured by ddPCR from the pure water field blank averaged 70 ± 9% (mean ± SD, n = 16) of the spiked eluate control.

#### Amount of spiked-in Phi6 recovered from wastewater varies from site to site

Over the same period during which the amount of Phi6 measured in the field blank was evaluated (15 November to 29 December 2021), we similarly spiked 10 mL wastewater samples with Phi6 and then purified RNA using the Chemagic™ procedure. The amount of spiked-in Phi6 measured in wastewater from 21 sewersheds, sampled on 7 - 10 occasions for each site, averaged 29 ± 20% of the Phi6 eluate control, significantly less than the 70 ± 9% that was measured in spiked-in field blank controls (p<0.001, t-test). The amount of spiked-in Phi6 measured in wastewater samples averaged 39 ± 18% of the spiked-in Phi6 measured in same day field blanks. For a subset of these wastewater samples we compared the amount of spiked-in Phi6 measured by ddPCR when the RNA had been purified by the Chemagic™ procedure versus RNA from the same set of wastewater samples that had been purified by the PEG/NaCl/Qiagen procedure. Chemagic™ samples averaged 25 ± 15% of the eluate blank (n= 10 wastewater samples); whereas, PEG/NaCl/Qiagen samples averaged 6.5 ± 3.3% of the comparable spiked-in Qiagen blank (p<0.01, paired t-test, two tailed).

While the Chemagic™ process clearly enabled the measurement of more of the spiked-in Phi6 than the PEG/NaCl/Qiagen method, Table 1 shows that a significant amount of variation occurred in the amount of spiked-in Phi6 measured in wastewater samples collected from different locations (One way ANOVA, p<0.0001). As percent of the eluate controls, the amount of spiked-in Phi6 in wastewater samples varied from as low as 5 ± 3% for wastewater samples from site WH to as high as 53 ± 13% for wastewater samples from site DB (significantly different, post-hoc Tukey, p<0.05).

To determine if the variation between Phi6 measured from different sites might be due to varying amounts of PCR inhibitors present in the purified RNA, these results are compared to a set of experiments in which the Phi6 was spiked into all wastewater samples at the elution step and compared to the elution control for the same experiments (Table 2). This procedure was done for 52 wastewater samples and 5 field blanks, yielding an average amount of Phi6, compared to Phi6 spiked into the eluate control, of 98.9 ± 9.6% for wastewater samples and 96.0 ± 5.4% for the field blanks. This indicates that, on average, PCR inhibition due to factors in the eluted RNA solution did not occur. Thus, the differences in Phi6 measurements observed for the various sites when Phi6 was spiked into wastewater before Chemagic™ processing were due to differences in recovery of RNA in the purification process, rather than the presence of PCR inhibitors in the eluted RNA solutions.

**Table 2.** Phi6 spiked into wastewater purified by Chemagic™, as percent of eluate control.

|  |  |  |  |  |
| --- | --- | --- | --- | --- |
| Sample | Average | SD | N | significance post-hoc Tukey Test* |
| Field blank | 70% | 9% | 16 | a |
| DB | 53% | 13% | 7 | a, b |
| KF | 51% | 18% | 7 | a, b, c |
| CT | 45% | 17% | 8 | b, c, d |
| BG | 42% | 5% | 7 | b, c, d, e |
| PA | 41% | 12% | 8 | b, c, d, e |
| CS | 39% | 19% | 8 | b, c, d, e |
| LW | 39% | 12% | 7 | b, c, d, e |
| HP | 37% | 13% | 7 | b, c, d, e |
| CC | 36% | 23% | 8 | b, c, d, e |
| ER | 34% | 16% | 7 | b, c, d, e, f |
| SG | 33% | 17% | 8 | b, c, d, e, f |
| SS | 21% | 5% | 7 | c, d, e, f |
| SE | 20% | 14% | 8 | d, e, f |
| EB | 20% | 9% | 7 | d, e, f |
| JV | 19% | 16% | 7 | d, e, f |
| HH | 18% | 8% | 7 | d, e, f |
| UC | 18% | 16% | 9 | d, e, f |
| ME | 17% | 13% | 8 | d, e, f |
| AS | 15% | 17% | 10 | e, f |
| WG | 14% | 16% | 7 | e, f |
| WH | 5% | 3% | 7 | f |

\*groups labeled with the same letters were not significantly different from one another at $p < 0.05$

**Table 3.** Phi6 spiked into Chemagic™ wastewater eluate as percent of eluate control.

|  |  |  |  |
| --- | --- | --- | --- |
| Sample | Average | SD | N |
| field blank | 96.0% | 5.4% | 5 |
| AS | 74.9% | 29.7% | 3 |
| BG | 115.5% | 16.3% | 3 |
| CC | 103.8% | 6.0% | 2 |
| CS | 104.0% | 4.8% | 2 |
| CT | 95.9% | 1.1% | 2 |
| DB | 89.6% | 9.4% | 5 |
| EB | 94.8% | 3.2% | 2 |
| ER | 106.1% | 2.7% | 2 |
| HH | 105.3% | 20.1% | 3 |
| HP | 99.8% | 20.0% | 3 |
| JV | 111.6% | 21.7% | 2 |
| KF | 97.6% | 6.3% | 2 |
| LW | 97.8% | 24.8% | 2 |
| ME | 99.2% | 3.8% | 2 |
| PA | 106.9% | 16.8% | 3 |
| SG | 98.3% | 3.7% | 2 |
| SS | 92.5% | 27.3% | 3 |
| UC | 93.1% | 15.9% | 3 |
| WG | 100.1% | 30.8% | 3 |
| WH | 91.3% | 11.6% | 3 |
| average of all sites | 98.9% | 9.6% | 20 sites |

#### More sensitive detection of SARS-CoV-2 with Chemagic™, compared to PEG/NaCl/Qiagen

The performance of Chemagic™ for detection of SARS-CoV-2 markers in wastewater was compared to the PEG/NaCl/Qiagen method. On several occasions during August, October, and November 2021 wastewater samples from various sewersheds in Detroit were processed with both the Chemagic™ and PEG/NaCl/Qiagen methods, spanning a period in which the markers were rarely detected by either method (August), only detected in about half the samples (October), and detected in most samples (November). Out of a total of 20 samples analyzed on the dates for which both methods were used, 9 samples occurred for which SARS-CoV-2 was detected in wastewater by one method and not the other. For all 9 samples, SARS-CoV-2 was detected in samples processed via the Chemagic™ method and not after processing by PEG/NaCl/Qiagen (significantly different, Fisher exact test, p < 0.0001). Figure 1 illustrates a representative example of the greater number of positive droplets detected after Chemagic™ purification than after PEG/NaCl/Qiagen purification.

**Figure 1:**
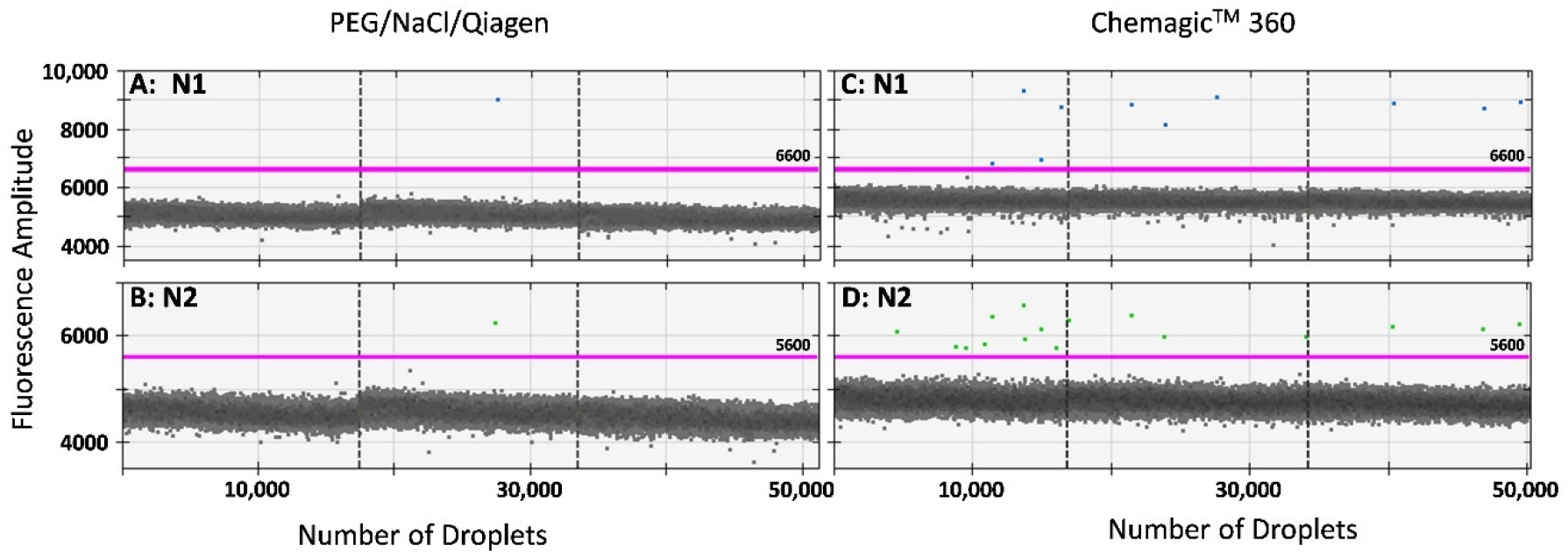
Representative example of detection of N1 and N2 from the same wastewater sample processed by Chemagic™ compared with PEG/NaCl/Qiagen, out of nine for which detection occurred with one method but not the other (all nine were positive for the Chemagic™ purified sample but not for PEG/NaCl/Qiagen; see text). ddPCR droplet results are shown for the same wastewater sample processed by PEG/NaCl/Qiagen (A: N1 and B: N2) and Chemagic™ (C: N1 and D: N2). All ddPCR results are technical triplicates with dashed vertical line separating the droplet results of the three reactions. Positive droplets are the points above the horizontal threshold line; the broad band of droplets below the threshold are negative droplets, approximately 17,000 per reaction. Bio-Rad considers three or more positive droplets to yield reliable Poisson statistics, which was achieved for this sample using Chemagic™ purification but not for PEG/NaCl/Qiagen.

#### Higher amount of SARS-CoV-2 measured in wastewater samples processed via Chemagic™ compared to the PEG/NaCl/Qiagen method

For wastewater samples for which both PEG/NaCl/Qiagen and Chemagic™ methods yielded detectable SARS-CoV-2 markers, the amount of SARS-CoV-2 measured by ddPCR was usually greater with the Chemagic™ method. In our data set, 10 sites processed by both methods had detectable signal from at least 1 of the 2 gene markers. The N1 gene target was detected in all 10 samples processed via Chemagic™ but only 8 of the samples processed by PEG/NaCl/Qiagen. The N2 gene target was detected in all 10 samples processed by Chemagic™ while only 9 samples processed by PEG/NaCl/Qiagen were detectable. The quantity of viral copies per 100 mL of wastewater for all samples that were above the limit of detection for each method was correlated, as illustrated in the linear regression in Figure 2. In all but 2 samples the Chemagic™ purification method produced more signal than its PEG/NaCl/Qiagen counterpart (significantly different for both markers: N1: Paired t-test, two-tailed p = 0.007; N2: Paired t-test, two-tailed p = 0.013). On average, the number of copies of SARS-CoV-2 detected per 100 mL in the Chemagic™-purified samples was 4.9 ± 3.4 times the number of copies measured in the PEG/NaCl/Qiagen-purified samples.

**Figure 2.**
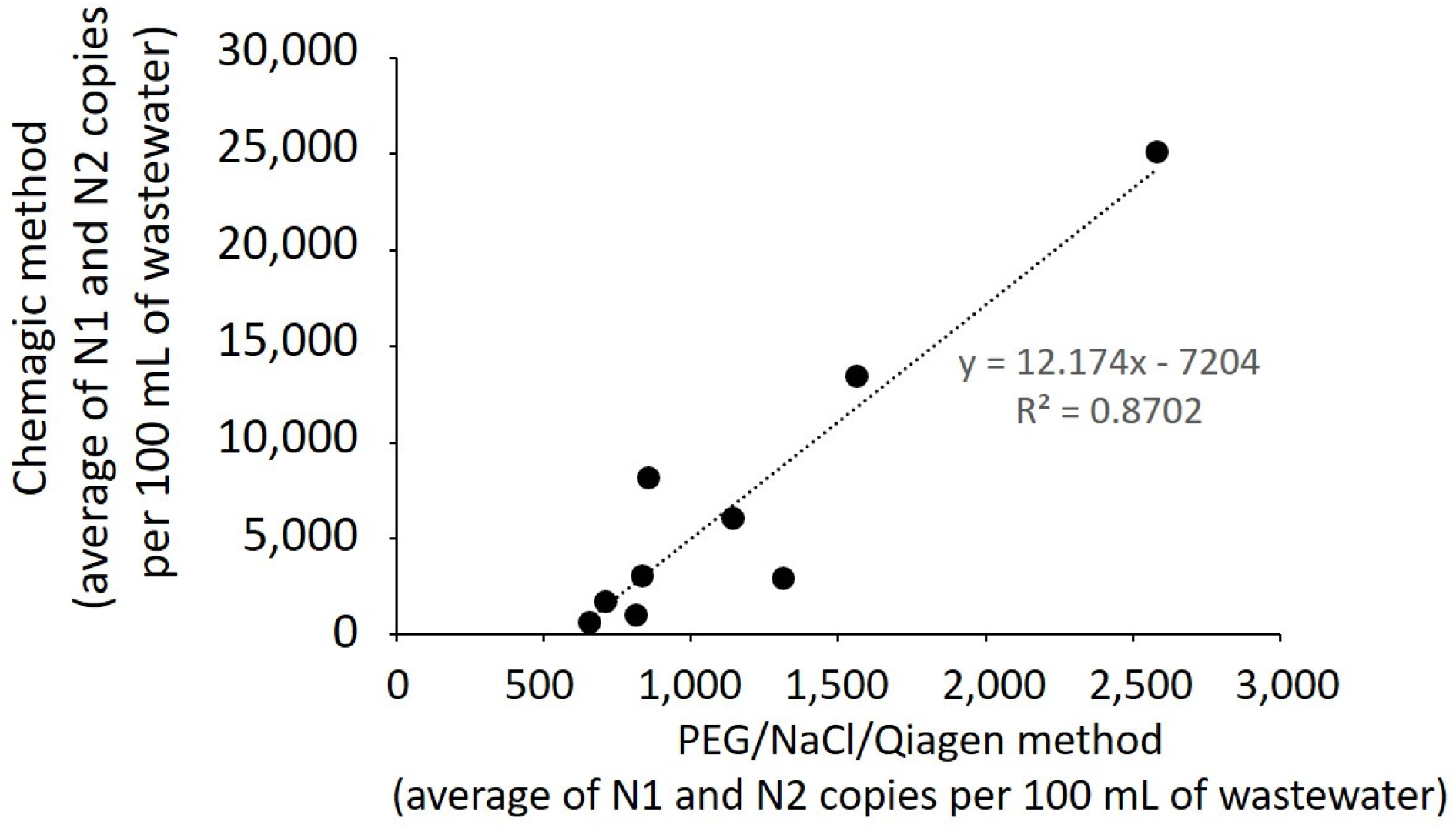
Comparison of the amounts of N1 and N2 markers measured in samples purified by the Chemagic™ method v the PEG/NaCl/Qiagen method, for samples in which both methods yielded a detectable measurement. Note the much higher values on the Chemagic™ axis v the PEG/NaCl/Qiagen axis.

#### Time course of SARS-CoV-2 detected in wastewater from three sites in Detroit

As demonstration of the application of wastewater analysis to small sewersheds in Detroit, we report here SARS-CoV-2 marker data for samples collected from 3 sites in Detroit from the second week of August through the last week of December. Initially, the PEG/NaCl/Qiagen method was used for sample processing and continued until the last week of September, after which samples were processed on the Chemagic™ platform. During August and September the majority of samples had no detectable signal and this trend continued after switching to Chemagic™. Beginning in November we observed increases in samples from all three sites, with two of them rising simultaneously to a high level (and the third one moderately) on the last illustrated sampling day, the beginning of the “omicron surge.”

## Discussion

This paper demonstrates here an automated purification procedure based on application of the 12-Rod Perkin-Elmer Chemagic™ 360 platform to detection of SARS-CoV-2 in wastewater. Compared to the PEG/NaCl/Qiagen method that we formerly used, the Chemagic™ system enabled a more sensitive detection of SARS-CoV-2 at the lowest detectable limit, as exemplified by the 9 wastewater samples in which we detected SARS-CoV-2 markers after purification with Chemagic™ but not after purification by PEG/NaCl/Qiagen. This greater sensitivity is further demonstrated in quantitative comparisons showing the recovery of 4-times as much Phi6 spiked-in RNA (25 ± 15% of the eluate control for Chemagic™ v 6.5 ± 3.3% for the PEG/NaCl/Qiagen method) and the 4.9:1 fold ratio of amount of N1 and N2 measured in same sample comparisons of the two methods (see text and Figure 2). Application of the Chemagic™ method enabled sensitive detection of the N1 and N2 markers in wastewater from three small sewersheds illustrated in Figure 3; these data are part of a larger city-wide project to monitor 20 sewersheds in Detroit, illustrated in a public wastewater dashboard at https://www.ramlabwsu.org/public-data-page.html.

**Figure 3.**
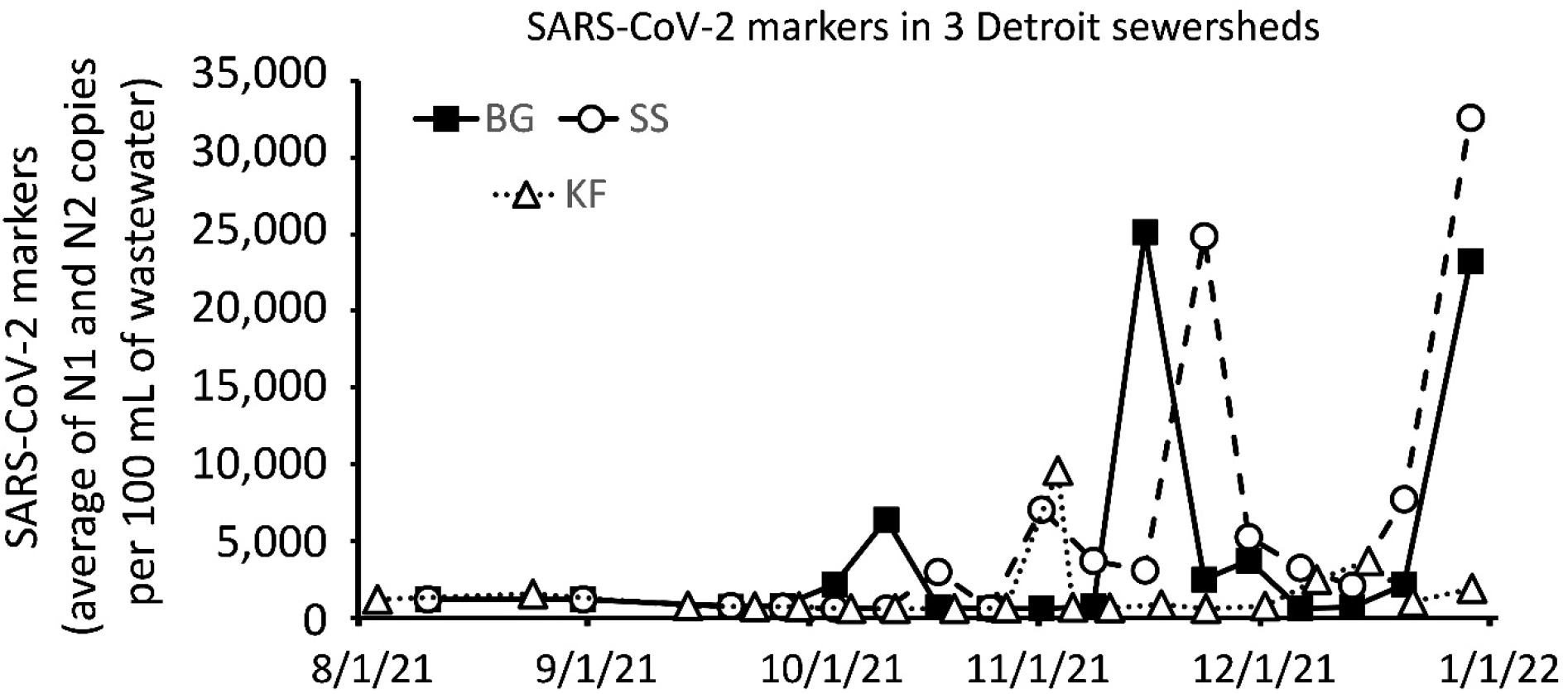
Levels of SARS-CoV-2 markers in wastewater collected from 3 sewersheds in Detroit located in southeast, central, and northwest areas of the City. These are small sewersheds with estimated populations ranging from 400 to 1,000 people and therefore their exact locations are restricted. On October 4, the method of viral RNA purification was changed from PEG/NaCl/Qiagen to Chemagic™ 360.

A previous paper demonstrating the application of a PEG/NaCl/Qiagen method in Michigan similar to the one used here, by Flood et al. (2021) reported obtaining a Phi6 recovery from the initial amount spiked into wastewater of about 20%, which is 3-fold higher than we report here for the PEG/NaCl/Qiagen method. The recovery calculated by Flood et al. is for the PEG/NaCl concentration step and does not include additional losses, if any, from the Qiagen purification. Using a comparable PEG/NaCl/RNA purification method, Torii et al. (2022) reported RNA virus recoveries of 0.07% - 2.6%, and values summarized from recent publications using PEG/NaCl/Qiagen (or other commercial RNA purification) methods, ranging from less than 1% to as high as 50%.

Further considerations for choosing a purification method for RNA from wastewater include expense of equipment, cost of reagents, staff time, and complexity or problems with the method. In choosing the Chemagic™ instrument for our laboratory, we tested and compared several instruments based on on-site demonstrations and descriptions provided by vendors. Methods tested included the PEG/NaCl/Qiagen method as described by the State of Michigan protocol based on the work of Flood et al. (2021), the Promega Maxwell® RSC system, and the PerkinElmer Chemagic™ 360. The Kingfisher by Thermofisher was evaluated by analysis of the protocols described in publications (Karthikeyan and Humphrey, 2020; Appliedbiosystems, 2021; Karthikeyan et al., 2021).

The PEG/NaCl/Qiagen method requires the availability of a large-volume refrigerated centrifuge. The large volume and lengthy time (2 hr) of the incubation step and (45 min) of the centrifugation step (100 mL in 250 mL centrifuge tubes) potentially limits the throughput of samples per day. Both the Promega protocol and the Chemagic™ protocol require an early centrifugation step to remove large particulates; however, the Chemagic™ centrifugations are relatively brief (15 min) and can be done in 50 mL conical tubes, enabling larger numbers of samples to be processed per unit time than the PEG/NaCl/Qiagen method.

The Promega protocol that we tested utilized a non-automated vacuum-operated Pure Yield Midi Column method to concentrate wastewater down to a volume of 1 mL, which was necessary for applying the sample to their Maxwell RSC automated instrument. This method can work well; however, we found that some wastewater samples clogged the columns despite the preceding centrifugation step whose purpose was to reduce the likelihood of clogging. In those runs, it so happened that the samples that clogged the columns were among the most “necessary,” in that they had been exhibiting higher viral levels than several other samples. Samples with fewer suspended solids worked well, but the potential loss of samples that we were most interested in was a problem for us. In contrast to the several pipetting and decanting steps in the non-automated part of the procedure, the automated part of the procedure with the Maxwell RSC instrument was easy to perform. We found that the design using cartridges pre-filled with reagents in the instrument eliminated a lot of pipetting steps that other systems required at various points.

The Kingfisher wastewater method has been described in a published protocol (Karthikeyan and Humphrey, 2020; Karthikeyan et al., 2021) and User Guide (Appliedbiosystems, 2021). While framed as a fully automated system, the protocols describe a large number of manual pipetting steps in the set-up of reagents into 9 plates (three 24-well plates and six 96-well plates) that are to be put into the instrument. In addition, the method requires two sequential automated procedures between which samples must be manually transferred from a 24-well format to a 96-well format.

The reagents for the Chemagic™ system are held in large reservoirs that serve for multiple runs (up to 250 samples before needing to be replenished). The Kingfisher has the flexibility that the types and concentrations can be more easily swapped or adjusted between runs (the user fills these in before each run) for optimizing purifications dependent on sample type; however, for purifications with identical reagents repetitively used run-to-run, the Chemagic™ is convenient to use.

We compared the cost of materials needed per sample to go from raw wastewater to PCR-ready RNA. These estimates were developed from a detailed review of the described methods and our direct experience testing demo models and our use of the PEG/NaCl/Qiagen method for 10 months, prior to beginning our regular use of the Chemagic™ instrument. The estimated costs include not only the reagents required by the method (least expensive for the PEG/NaCl/Qiagen method) but also the plastics (tips, tubes) and personal protective equipment that was necessary and required by our institution for the various methods. Rounded to the nearest $5 per sample, the cost of supplies per sample was $20 for PEG/NaCl/Qiagen, $30 for Chemagic™ 360, and $40 for Promega Maxwell and Kingfisher. These costs do not take into account the differences in time and labor; however, based on our experience, the labor involved in using the Chemagic™ 360 is much less than for the PEG/NaCl/Qiagen method.

Estimating the cost of the equipment depends somewhat on how much one might value or need to spend additionally on accessories, such as stir manifolds and refrigeration for the PEG/NaCl/Qiagen method, vacuum pump for the Promega column concentration method, hotplates required for some methods, and the type and speed of centrifuges that might be required. Disregarding those additional costs, we found that the Promega Maxwell RSC instrument was the least expensive automated system and the Chemagic™ 360 instrument was the highest-priced.

We chose the Chemagic™ instrument on the basis of lower labor requirements and the idea that for the number of samples we plan to process, the cost-savings for Chemagic™ supplies compared to the other automated instruments would make up the difference in instrument cost within two years of owning the instrument. While the PEG/NaCl/Qiagen method is considerably less expensive both in equipment requirements and supplies, the faster processing time and greater sensitivity of SARS-CoV-2 detection compared to the PEG/NaCl/Qiagen method is important to obtain the earliest warning of resurgences of infections in the community.

## Data Availability

All data produced in the present study are available upon reasonable request to the authors, except that the exact locations of the three small sewersheds for which data are provided in Figure 3 is restricted and cannot be revealed without permission of the Detroit Health Department.

https://www.ramlabwsu.org/public-data-page.html

https://dx.doi.org/10.17504/protocols.io.b2reqd3e

## Declaration of Competing Interest

The authors have no known competing personal relationships or financial interests that could have influenced the work reported in this paper.

## Acknowledgements

Funding for this work was provided by Project AY of the WSU-MDHHS Master contract MA-2021, entitled “SARS-CoV-2 Epidemiology – Wastewater Evaluation and Reporting (SEWER) Network” (Principal Investigators: Jeffrey L. Ram and William Shuster) from the Michigan Department of Health and Human Services, using Federal Financial Assistance from the U.S. Department of Treasury under the Epidemiology and Laboratory Capacity: Enhancing Detection Expansion through Coronavirus Response and Relief (CRR) Supplemental Appropriations Act of 2021 (P.L. 116-260). Wastewater samples were collected with cooperation and guidance from the Detroit Water and Sewerage Department, the Detroit Health Department, the Wayne State University Campus Health Center, and the Great Lakes Water Authority.

## Notes

### Competing Interest Statement

The authors have declared no competing interest.

### Funding Statement

This study was funded by Project AY of the WSU-MDHHS Master contract MA-2021, entitled "SARS-CoV-2 Epidemiology: Wastewater Evaluation and Reporting (SEWER) Network" (Principal Investigators: Jeffrey L. Ram and William Shuster) from the Michigan Department of Health and Human Services, using Federal Financial Assistance from the U.S. Department of Treasury under the Epidemiology and Laboratory Capacity: Enhancing Detection Expansion through Coronavirus Response and Relief (CRR) Supplemental Appropriations Act of 2021 (P.L. 116-260).

### Author Declarations

The IRB Administration office of Wayne State University waived ethical approval for this work in WSU IRB HPR number 2021 190.

